# An open-label, parallel-group, randomized clinical trial of different silver diamine fluoride application intervals to arrest dental caries

**DOI:** 10.1101/2024.03.26.24304906

**Authors:** Robert J. Schroth, Sukeerat Bajwa, Victor H. K. Lee, Betty-Anne Mittermuller, Sarbjeet Singh, Vivianne Cruz de Jesus, Mary Bertone, Prashen Chelikani

## Abstract

**Background:** Non-surgical interventions are preferred to address the widespread issue of early childhood caries (ECC). Silver diamine fluoride (SDF) is an antimicrobial agent and alternative treatment option that can be used to arrest dental decay. While there is optimism with SDF with regard to caries management, there is no true consensus on the number and frequency of applications for children. The purpose of this study was to examine the effectiveness of 38% SDF to arrest ECC at three different application regimen intervals.

**Methods:** Children with ECC were recruited from community dental clinics into an open-label, parallel-group, randomized clinical trial. Participants were randomized to one of three groups: visits one month, four months, or six months apart. Participants received applications of 38% SDF, along with 5% sodium fluoride varnish (NaFV), at the first two visits to treat cavitated carious lesions. Lesions were followed and arrest rates were calculated. Lesions were considered arrested if they were hard on probing and black in colour. Statistics included descriptive and bivariate analyses. A *p*-value of ≤ 0.05 was considered significant.

**Results:** Eighty-four children participated in the study (49 males and 35 females, mean age: 44.4 ± 14.2 months). Treatment groups were well matched with 28 participants per group. A total of 374 teeth and 505 lesions were followed. Posterior lesions represented only 29.1% of affected surfaces. Almost all SDF treated lesions were arrested for the one-month (98%) and four-month (95.8%) interval groups at the final visit. The six-month group experienced the lowest arrest rates; only 72% of lesions were arrested (*p* < 0.001). The duration of application intervals was inversely associated with improvements in arrest rates for all lesions.

**Conclusions:** Two applications of 38% SDF and 5% NaFV in one-month and four-month intervals were comparable and very effective in arresting ECC. Applications six months apart were less effective and could be considered inferior treatment.

**Trial registration:** ClinicalTrials.gov NCT04054635 (first registered 13/08/2019).

## INTRODUCTION

Early childhood caries (ECC), defined as the presence of dental caries in the primary dentition of children under six years of age, is a significant issue. Recent prevalence estimates in Canada range from 28% to 98% (1–3). The American Academy of Pediatric Dentistry (AAPD) recognizes the widespread and virulent nature of ECC, and supports the implementation of non-surgical interventions whenever possible (4). Non-surgical interventions delay or decrease the need for dental surgery to treat severe cases of ECC. Conscious sedation or general anesthesia in operating rooms are frequently used to facilitate restorative treatment of young children with ECC. However, they come with increased costs for treatment and greater risks for the child. Restorative treatment is still the predominant method of managing ECC. It is important to note that restorative treatment alone does not address the underlying cause of ECC. Consequently, there is a high risk of recurrence and many children form new carious lesions (5, 6).

Unfortunately, many children experience limited access to dental care and go through life with untreated caries, which can pose a serious health risk (7). The consequences of ECC are comprehensive. They include greater risk of carious lesion in the primary and permanent dentition, increased hospitalization and emergency visits, higher treatment costs, and reduced oral health-related quality of life (5, 8, 9). Furthermore, ECC can affect a child’s nutritional status and disrupt school attendance and performance (10–14). The multifactorial nature of ECC creates challenges in identifying effective primary prevention strategies (15). There were no effective non-surgical products available for secondary prevention until recently.

Reports have identified silver diamine fluoride (SDF) as an antimicrobial agent that can successfully arrest dental decay (16). It can potentially address untreated caries in young children, which would reduce the need for rehabilitative dental surgery under general anesthesia (17–22). SDF is a good alternative for children with ECC who may not be cooperative with traditional treatment approaches (23, 24). One systematic review with meta-analysis found that SDF was safe and effective in arresting dental caries in primary teeth. In eight studies that used 38% SDF to treat active caries, 81% of lesions were arrested (25). The American Dental Association (ADA) practice guidelines for non-restorative treatments of dental caries recommends the prioritization of 38% SDF over other products to manage cavitated carious lesions (26). Despite this information, true consensus on the frequency of SDF applications for children with ECC is lacking. The current AAPD clinical practice guidelines for SDF urge researchers to conduct well-designed randomized clinical trials to compare the use and outcomes of SDF treatment on both primary and permanent teeth (27).

While Advantage Arrest^TM^ (38% SDF) received approval for clinical use in Canada in 2017, there has been little guidance on the frequency and duration of its application. Proposed protocols may not translate well into some clinical and dental public health settings. Recommendations for frequent re-application may not be practical or realistic in remote communities where access to dental care is limited and where frequent follow-up visits are not possible in a short amount of time (20, 25, 28).

The purpose of this study was to examine the effectiveness of SDF to arrest cavitated carious lesions in primary teeth at three different application regimen intervals (one month, four months, and six months apart). To our knowledge, this is the first randomized clinical trial of SDF conducted in Canada for young children. This study aimed to provide new information that may aid clinicians in the decision-making process for SDF application for the greater benefit of patients.

## METHODS

This open-label, parallel-group, randomized clinical trial was registered at ClincialTrials.gov (registration number: NCT04054635, first registered 13/08/2019). Participants were recruited between October 2019 and June 2021 from community dental clinics in Winnipeg, Canada (Access Downtown, Mount Carmel Clinic, and SMILE plus). Study visits also took place at the Children’s Hospital Research Institute of Manitoba. Children under 72 months of age were included if they had teeth that met International Caries Detection and Assessment System (ICDAS) codes 5 or 6 criteria, with softer caries extending into dentin without signs of pulpal involvement (29). Children were excluded if they had a silver allergy, developmental enamel defects, severe medical issues, dental conditions requiring immediate rehabilitation under general anesthesia, or if they had teeth that met any PUFA (Pulpal involvement, Ulceration, Fistula, and Abscess) index criteria. Analyses of radiographs were not conducted, as not every child had them done. Parents/caregivers provided written informed consent.

A total of 84 participants were recruited for the study. Sample size was determined based on a pilot study and in consultation with a biostatistician. In the pilot study, 40 children had 239 lesions (approximately six lesions per child) that could estimate an arrest rate with a 95% confidence interval (CI) to be accurate within ± 6.5%. With at least 400 lesions in a proposed sample, the 95% CI would be ± 5%. Anticipating an average of six lesions per child, three regimen groups with 23 children each would produce approximately 414 lesions to be studied. To deal with potential drop-outs/loss to follow-up, we over-recruited by 27.3% and sought 28 children for each group.

Participants came for three study visits (Figure 1). Children underwent dental examinations at each visit. Teeth meeting ICDAS codes 5 or 6 criteria were identified at baseline, and the location, size, hardness (soft, medium, or hard), colour (yellow, brown, or black), and activity of lesions were recorded. Dmft (decayed, missing, and filled primary teeth) index scores were calculated. Lesions were treated with 38% SDF (Advantage Arrest, Oral Science, Brossard, Québec, Canada) at the first and second visits and were followed for the duration of the study. The liquid product was applied with a microbrush for one minute, and surfaces were wiped with wet gauze and rinsed with water. Participants received applications of 5% sodium fluoride varnish (NaFV) following SDF application. One attending dentist carried out all clinical activities, while other research staff conducted all non-clinical activities. Parents/caregivers were also administered questionnaires at each visit. The questionnaires asked for information on sociodemographic characteristics, oral hygiene, pain, oral health-related quality of life, and the appearance of teeth.

**Figure 1.**
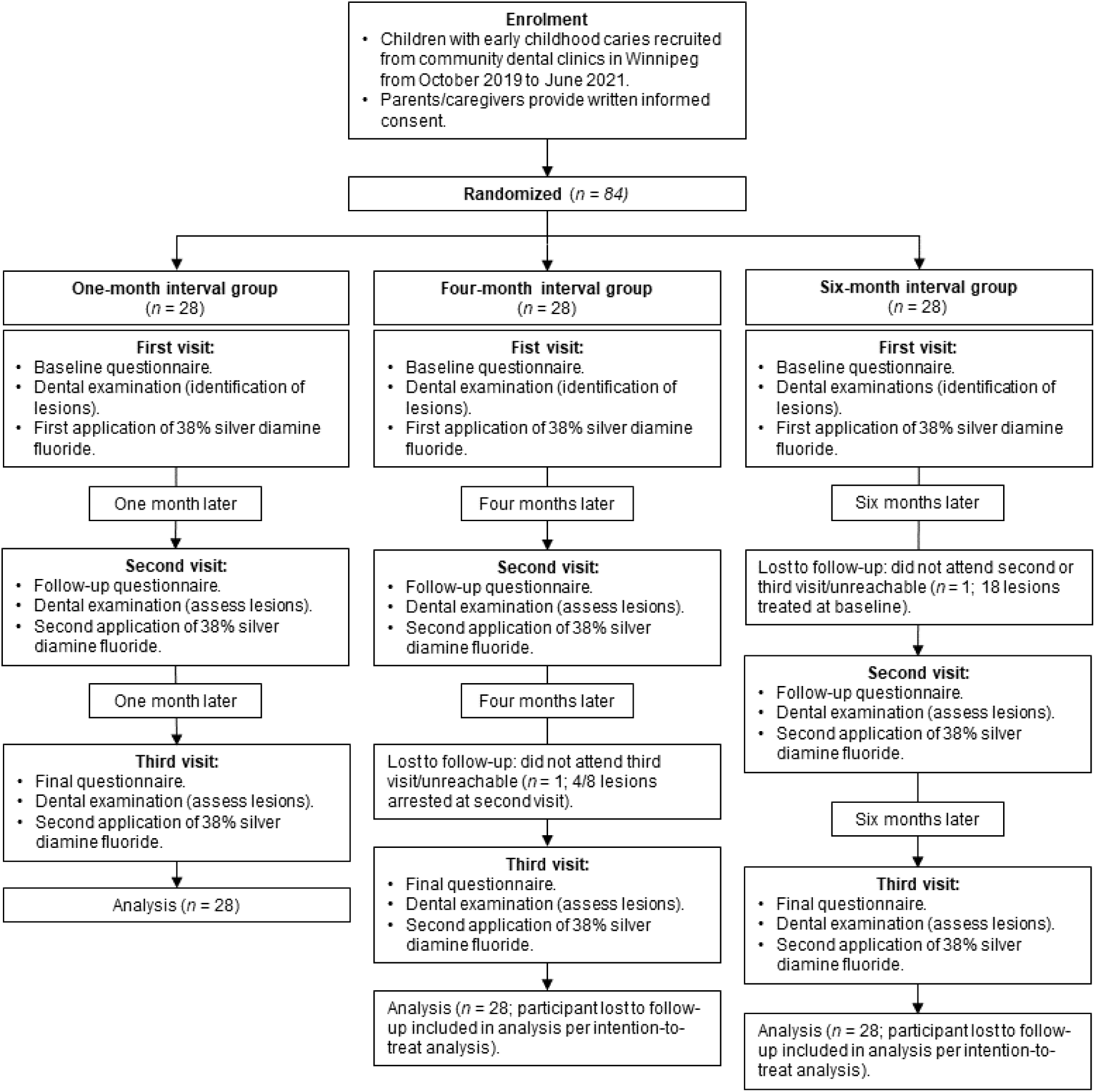
Flow diagram of study process (recruitment, randomization, visits and activities, duration, and analysis)

The time between SDF treatments and study visits depended on the child’s regimen. Prior to recruitment, the research coordinator prepared sealed envelopes containing details for one of three regimens: treatment/visits one month apart (proposed in the AAPD’s clinical practice guideline), four months apart (protocol frequency adopted by the Winnipeg Regional Health Authority), or six months apart (recommended by ADA) (30–32). When a child was recruited into the study, research staff randomly selected an envelope, thus assigning the child to one of the three groups. Participants were followed for a total of two months, eight months, or 12 months. The first participant was enrolled 19 October 2019, and the last participant was seen 12 February 2022. Examiners and research staff were not blinded to the prior status of lesions.

The primary outcome measure was arrest rates among individual treatment groups. Lesions that were hard upon tactile probing and black in colour were considered arrested. Overall arrest rates and specific arrest rates for anterior (primary incisors and canines) and posterior (primary molars) lesions at the second and third visits were calculated. Intention-to-treat analysis was used, where participants lost to follow-up were still included in the study, and we acted as though there were no changes to lesions for these individuals at subsequent (missed) visits. This approach was chosen since it preserved randomization and was the best neutral response for the unknown status of lesions—to assume no effect either way (33). Data were entered into REDCap (Research Electronic Data Capture), a secure web application for online databases, and were analyzed using Number Cruncher Statistical Software Version 9.0 (NCSS; Kaysville, Utah). Descriptive statistics were also calculated for relevant questionnaire information. Kruskal-Wallis one-way analysis of variance (ANOVA) and Pearson’s Chi-squared test were performed when appropriate. A *p*-value of ≤ 0.05 was considered statistically significant.

The ADA maintains that 5% NaFV is largely unproductive as a treatment for cavitated lesions (26). We did not consider a control group receiving only 5% NaFV, as this would be considered unethical substandard care. Sodium fluoride is included in treatments following the application of SDF because it prevents caries on surfaces by strengthening the tooth structure and increasing resistance to acidic demineralization (30, 34).

## RESULTS

Participant characteristics are summarized in Table 1. Forty-nine male participants and 35 female participants were randomized into three groups of 28 children. The mean age of children recruited into the study was 44.4 ± 14.2 months. The overall sample was diverse, with participants having different African (38.1%), Asian (28.6%), European (9.5%), or Canadian Indigenous (23.8%) ancestry. Few children (16.7%) were newcomers to Canada. There were no significant differences between the three groups in terms of age, sex, and ethnicity. A majority of participants brushed their teeth twice a day (61.9%) and used toothpaste containing fluoride (82.1%). Most participants (69.1%) also had some form of dental insurance that covered all or part of their dental care expenses. These results were consistent across all three groups in the study. Only five children experienced any tooth pain at their first study visit.

**Table 1.**
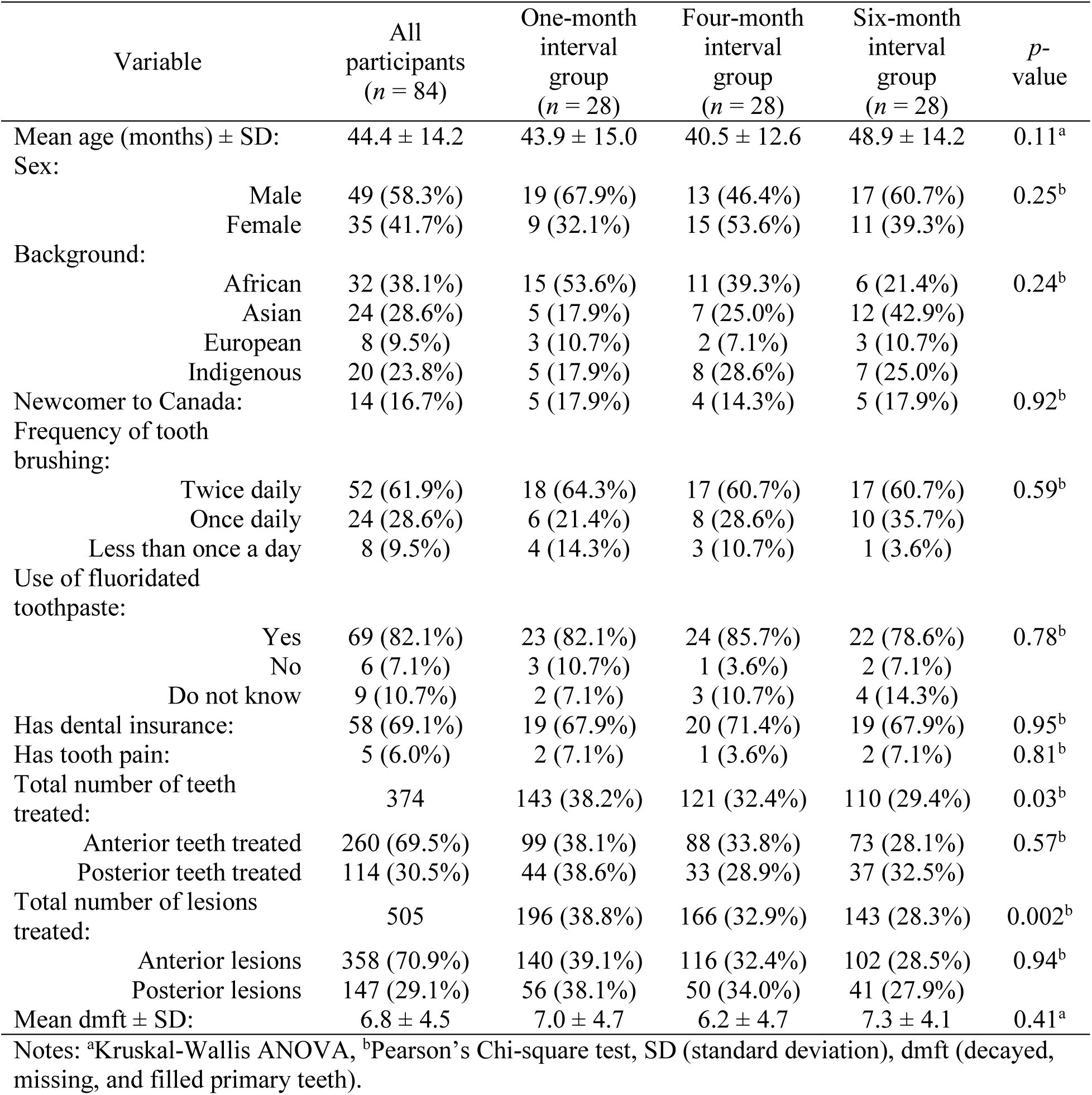
Participant characteristics at baseline.

Two participants were lost to follow-up. A child in the four-month interval group did not attend their third visit (4/8 lesions arrested at second visit; 2/2 anterior lesions and 2/6 posterior lesions), and a child in the six-month group did not attend either of their follow-up visits (18 lesions treated at baseline; 12 anterior lesions and six posterior lesions). Because of intention-to-treat analysis, we assumed no changes in lesion status for these children since their last visit (i.e., 4/8 lesions were recorded as arrested at the third visit for the child in the four-month group, and no lesions were recorded as arrested at subsequent visits for the child in the six-month group).

A total of 374 teeth and 505 lesions were treated with 38% SDF and 5% NaFV. The number of teeth differed significantly by group classification (*p* = 0.03), with 143 teeth treated in the one-month interval group, 121 teeth treated in the four-month interval group, and 110 teeth treated in the six-month interval group. The number of lesions also differed significantly by group classification (*p* = 0.002); the one-month interval group had 196 lesions treated, the four-month interval group had 166 lesions treated, and the six-month interval group had 143 lesions treated. More anterior teeth (260) and lesions (358) were treated than posterior teeth (114) and lesions (147). The number of anterior and posterior teeth and lesions did not vary significantly between groups. Kruskal-Wallis one-way ANOVA found no significant difference between groups in mean dmft. Overall, participants had a mean dmft of 6.8 ± 4.5.

Lesion arrest rates are summarized in Table 2 and Figure 2. The one-month interval group and the four-month interval group had high arrest rates at the first follow-up after the initial application of SDF and NaFV, with 78.1% and 81.3% of lesions arrested, respectively. The six-month interval group had just 61.5% of lesions arrested at that time. At the second follow-up visit (i.e., the third and final visit), almost all lesions were arrested for the one-month (98%) and four-month (95.8%) interval groups. The six-month interval group only had 72% of lesions arrested at that time. Pearson’s Chi-squared test revealed significant associations between group classification and arrest rates (*p* < 0.001). The duration of the application regimen interval was inversely associated with improvements in arrest rates from the second to third study visit. The one-month interval group showed the greatest improvement in their condition with a 19.9% increase in arrested lesions, the four-month interval group was second with a 14.5% increase, and the six-month interval group showed the least improvement with a 10.5% increase.

**Figure 2.**
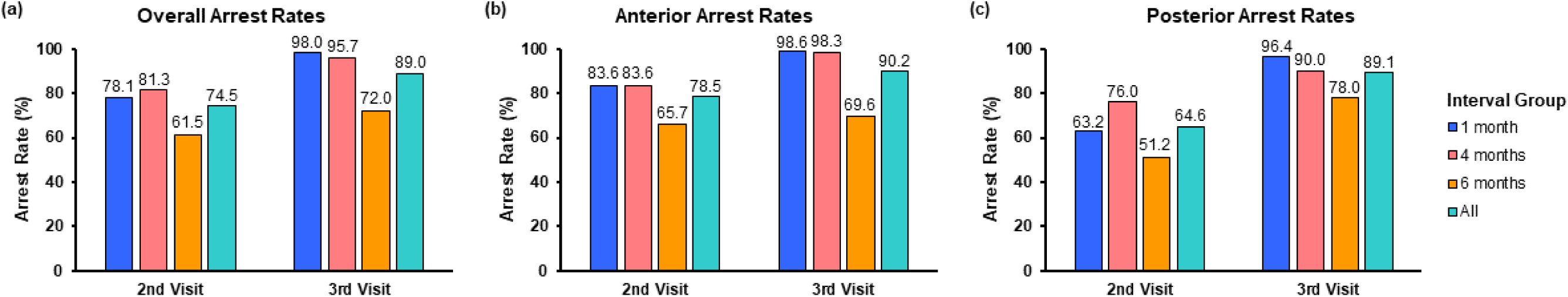
Early childhood caries arrest rates after SDF treatment for different application regimen intervals: (a) overall lesions, (b) anterior lesions, and (c) posterior lesions.

**Table 2.** Arrest rates after SDF and 5% NaFV application(s).

| Overall lesions arrest rates |  |  |  |
| --- | --- | --- | --- |
| Participants | At second visit<br>(first follow-up) | At third visit<br>(second follow-up) | Second to third visit<br>differential |
| All participants | 74.5% (376/505) | 89.9% (454/505) | +15.4% |
| One-month interval group | 78.1% (153/196) | 98.0% (192/196) | +19.9% |
| Four-month interval group | 81.3% (135/166) | 95.8% (159/166) | +14.5% |
| Six-month interval group | 61.5% (88/143) | 72.0% (103/143) | +10.5% |
| <sup>a</sup>*p* value | < 0.001 | < 0.001 |  |
| Anterior lesions arrest rates |  |  |  |
| Participants | At second visit<br>(first follow-up) | At third visit<br>(second follow-up) | Second to third visit<br>differential |
| All participants | 78.5% (281/358) | 90.2% (323/358) | +11.7% |
| One-month interval group | 83.6% (117/140) | 98.6% (138/140) | +15.0% |
| Four-month interval group | 83.6% (97/116) | 98.3% (114/116) | +14.7% |
| Six-month interval group | 65.7% (67/102) | 69.6% (71/102) | +3.9% |
| <sup>a</sup>*p* value | 0.001 | < 0.001 |  |
| Posterior lesions arrest rates |  |  |  |
| Participants | At second visit<br>(first follow-up) | At third visit<br>(second follow-up) | Second to third visit<br>differential |
| All participants | 64.6% (95/147) | 89.1% (131/147) | +24.5% |
| One-month interval group | 63.2% (36/56) | 96.4% (54/56) | +33.2% |
| Four-month interval group | 76.0% (38/50) | 90.0% (45/50) | +14.0% |
| Six-month interval group | 51.2% (21/41) | 78.0% (32/41) | +26.8% |
| <sup>a</sup>*p* value | 0.05 | 0.02 |  |

Note: <sup>a</sup>Pearson's Chi-square test.

Anterior-specific analyses showed higher arrest rates for primary incisors and canines with the one-month (83.6%), four-month (83.6%), and six-month (65.7%) interval groups at the first follow-up. Pearson’s Chi-squared test results were significant for these findings (*p* = 0.001). At the second follow-up visit, almost all lesions were arrested for the one-month (98.6%, +15.0% improvement) and four-month (98.3%, +14.7% improvement) interval groups. The six-month interval group, however, experienced less success and only had 69.6% of lesions arrested at that last visit (+3.9% improvement). These findings were significant (*p* < 0.001).

Posterior-specific arrest rates at the first follow-up for the one-month (63.2%), four-month (76.0%), and six-month (51.2%) interval groups were lower than overall and anterior-specific arrest rates at that time (*p* = 0.05). Almost all molar lesions were arrested for the one-month (96.4%) and four-month (90.0%) interval groups at the second follow-up visit. The six-month interval group only had 78% of lesions arrested at that time. All posterior-specific findings were significant (*p* = 0.02). Despite the low arrest rate at the first follow-up visit, the one-month interval group showed good improvement in their condition and had a 33.2% increase in arrested lesions. The six-month group also recovered and had a 26.8% increase in arrested lesions. The four-month group experienced a +14.0% differential.

## DISCUSSION

This randomized clinical trial investigated whether three different application intervals of 38% SDF, along with 5% NaFV, performed similarly with respect to arresting caries lesions. Overall, two applications of SDF and NaFV either one month or four months apart were very successful in arresting lesions in primary teeth and resulted in similar arrest rates. Applications six months apart were less successful and more lesions were not arrested. Shorter intervals between treatments (i.e., one month and four months) appeared to be more effective than longer intervals (i.e., six months). Greater improvements in conditions following primary applications of SDF and NaFV were seen for individuals with more immediate follow-up visits.

Research on the use of SDF is mixed and there is no consensus on the number or frequency of applications to arrest dental caries in children. Some studies have also shown underwhelming results with semi-annual applications of SDF. Mabangkhru et al. examined the results of 38% SDF applications in children at six-month intervals, and found low arrest rates at first (20.5%, 228/1111 lesions) and second (35.7%, 397/1111 lesions) follow-up visits. These results were greater than those seen in a 5% NaFV control group at first (12.3%, 140/1138 lesions) and second (20.9%, 238/1138 lesions) follow-up visits (*p* < 0.001) (35). Fung et al. repeated applications of SDF every six months for young children with ECC in Hong Kong, and found a comparable arrest rate of 75.7% (685/905 lesions) at a 30-month follow-up (23). Despite an increase in the amount of applications over a prolonged time frame, there was no outstanding difference in the outcome.

Conversely, additional time made a difference in a study conducted by Zhi et al., where semi-annual applications of 38% SDF became more effective over a two-year period. They found that arrest rates increased for each follow-up visit at six months (43.3%), 12 months (53%), 18 months (82.9%), and 24 months (90.7%) (36). In this case, treatments at six-month intervals worked with greater use of SDF and a longer wait. Our methods were more confined in this present study. Meta-analysis of data from eight clinical studies of SDF pooled results from six-month follow-ups and found that 86% of caries had arrested at that time (25). This is more optimistic than what we found.

Several studies have reported good success (arrest rates) with SDF (22, 31, 36). In some cases, SDF has worked quickly in treated lesions in primary teeth. Despite equivocal evidence, SDF is a valuable treatment option for dental caries in clinical and community settings (37). The adoption of an SDF intervention protocol has been shown to significantly reduce preventable dental hospitalizations, arrest caries in children that are unable to tolerate other restorative treatments, and improve oral health-related quality of life (38). Our study supports two applications of 38% SDF in one-month or four-month intervals to treat dental caries in children under 72 months of age. Two applications of SDF six months apart may be inferior treatment. Since the one-month and four-month groups were similar, our findings will undoubtedly be welcome news for busy dental public health programs and clinics in rural and remote regions where it may be next to impossible to have children return for re-application of SDF within a month of the initial application.

To our knowledge, this is the first randomized clinical trial of SDF conducted in Canada for young children. This study provides new information that may aid clinicians in the decision-making process for SDF application for the greater benefit of patients. Our sample of children recruited from community dental clinics in Winnipeg are representative of the target population with dental decay that requires SDF treatment. Hence, our results should be relevant to other considerations. These findings may have broader applications to other populations as well. The specificity of the inclusion criteria actually helps mitigate sampling bias, and similarities between treatment groups justify their comparability and allow us to interpret the relationship between intervention and outcome.

Our results are contingent on the use of intention-to-treat analysis, which attempts to be realistic in its assessment of an intervention (39). This approach preserves randomization and usually allows users to draw unbiased conclusions regarding the effectiveness of treatments (33). That said, it is important to note that 18 lesions were deemed not arrested for the one child in the six-month interval group that did not attend either of their follow-up visits. This number of lesions entails some ambiguity. An extremely optimistic view of SDF treatment could have involved a 12.6% increase in the overall arrest rate for the six-month regimen. However, even if all 18 of those lesions had been arrested, the percentage of successful treatments would still be lower than the one-month and four-month groups.

Another limitation of this study is the significant difference in the number of total teeth and lesions treated between the three groups. The number of teeth and lesions treated were in decreasing order from the one-month group, to the four-month group, and to the six-month group. Since arrest rates were analyzed using a pooled sample, the six-month interval group, along with the four-month interval group, may have been disadvantaged from the lack of additional teeth and lesions to be examined. Furthermore, anterior teeth have been shown to have higher arrest rates than posterior teeth when treated with SDF (24, 36, 40). Despite the comparability of the location of affected teeth between our groups, the disparity in anterior and posterior lesions may misrepresent the average effect of treatment. A greater number of posterior lesions could have been beneficial. It is also important to recognize the possibility of other unmeasured confounding factors that may have caused variation among participants and the outcomes of associated teeth/lesions.

## CONCLUSIONS

Two applications of 38% SDF, along with 5% NaFV, in one-month and four-month intervals were more effective in arresting ECC than two applications in six-month intervals. Findings from this study will help inform the refinement of existing clinical treatment protocols for SDF for use in dental public health settings. More clinical trials are needed to confirm the number and frequency of SDF applications to arrest caries lesions in young children.

## Data Availability

The datasets used and analyzed during the current study are available from the corresponding author on reasonable request.

## DECLARATIONS

### Ethics approval and consent to participate

This study was approved by the University of Manitoba’s Biomedical Research Ethics Board (HS22998/B2019:068). Parents/caregivers of child participants provided written informed consent.

### Consent for publication

Not applicable.

### Competing interests

The authors declare that they have no competing interests.

### Funding

Funds for this study were provided by the Children’s Hospital Research Institute of Manitoba and the Dr. Gerald Niznick College of Dentistry. Dr. Robert J. Schroth also held a Canadian Institutes of Health Research Embedded Clinician Researcher Salary Award.

### Authors’ contributions

RJS contributed to conception and design, data acquisition, analysis, and interpretation, drafting the manuscript, and critically revising the manuscript. SB contributed to design, data analysis, and interpretation, drafting the manuscript, and critically revising the manuscript. VHKL contributed to data acquisition, analysis, and interpretation, drafting the manuscript, and critically revising the manuscript. BAM contributed to data acquisition and analysis, and critically revising the manuscript. SS contributed to data acquisition and critically revising the manuscript. VCJ contributed to data acquisition, analysis, and interpretation and critically revising the manuscript. MB contributed to data interpretation and critically revising the manuscript. PC contributed to conception and design, data analysis and interpretation, and critically revising the manuscript. All authors read and approved the final manuscript.

## Acknowledgments

We would like to thank the staff at Access Downtown, Mount Carmel Clinic, and SMILE plus for their assistance with this study.

## LIST OF ABBREVIATIONS

AAPD: American Academy of Pediatric Dentistry
ADA: American Dental Association
ANOVA: analysis of variance
CI: confidence interval
dmft: decayed, missing, and filled primary teeth
ECC: early childhood caries
ICDAS: International Caries Detection and Assessment System
NaFV: sodium fluoride
NCSS: Number Cruncher Statistical Software
PUFA: Pulpal involvement, Ulceration, Fistula, and Abscess
REDCap: Research Electronic Data Capture
SD: standard deviation
SDF: silver diamine fluoride

## Notes

### Competing Interest Statement

The authors have declared no competing interest.

### Clinical Trial

NCT04054635

### Funding Statement

This study was funded by the Children’s Hospital Research Institute of Manitoba and the Dr. Gerald Niznick College of Dentistry. Dr. Robert J. Schroth also held a Canadian Institutes of Health Research Embedded Clinician Researcher Salary Award.

### Author Declarations

The University of Manitoba’s Biomedical Research Ethics Board gave ethical approval for this work (HS22998/B2019:068)

